# Minimally Important Difference of the FACE-Q Skin Cancer Module: A Distribution-Based and Anchor-Based Analysis

**DOI:** 10.64898/2026.02.12.26345803

**Authors:** Maarten J. Ottenhof

**Author notes:** [Co-author], [Senior Author]. **Corresponding Author:** Maarten J. Ottenhof, MD.

## Abstract

**Background:** The FACE-Q Skin Cancer Module is a validated patient-reported outcome measure for facial skin cancer surgery. However, the minimally important difference (MID)— the smallest change in score perceived as meaningful by patients—has not been established. Without the MID, individual score changes cannot be interpreted clinically. This study aimed to determine the MID for all four FACE-Q Skin Cancer scales.

**Methods:** Prospective cohort study at a tertiary center (2017–2018). Patients completed the FACE-Q preoperatively and at 1 week, 3 months, and 1 year postoperatively. MID was estimated using distribution-based methods (0.5 standard deviation, standard error of measurement) and an anchor-based approach using the FACE-Q Adverse Effects scale as an implicit anchor. Internal consistency (Cronbach’s α), effect sizes, and standardized response means were calculated.

**Results:** Of 287 enrolled patients, 111 had paired baseline–three-month data. All scales had strong internal consistency (α = 0.82–0.93). Cancer worry showed the largest improvement from baseline to three months (mean change −3.1 ± 5.8; SRM = −0.54; p < 0.001). When we combined the estimates, the MID values (sum scores / 0–100 scale) were: Appearance Satisfaction 2.0 / 5.6, Psychosocial Distress 2.0 / 6.2, Cancer Worry 2.5 / 6.2, and Scar Satisfaction 2.0 / 6.2. Anchor-based estimates for the Scar scale (2.4 sum points) confirmed distribution-based findings.

**Conclusions:** This study establishes the first MID values for the FACE-Q Skin Cancer Module. A change of approximately 2–2.5 sum points (5–6 points on a 0–100 scale) represents a minimally important difference across all scales. These thresholds enable clinicians and researchers to interpret individual FACE-Q score changes and design adequately powered clinical trials.

## INTRODUCTION

Patient-reported outcome measures (PROMs) have become essential endpoints in surgical research and clinical practice. The FACE-Q Skin Cancer Module is a rigorously developed, condition-specific PROM designed to evaluate outcomes in patients undergoing facial skin cancer surgery. It comprises four scales measuring appearance satisfaction, psychosocial distress, cancer worry, and scar satisfaction, each providing scores that reflect the patient’s subjective experience.

While the FACE-Q Skin Cancer Module has been translated, validated, and calibrated using modern psychometric methods including Rasch analysis and computerized adaptive testing, we still lack one key piece: the MID: the minimally important difference (MID) has not been established for any of its scales. The MID is defined as the smallest change in a PROM score that patients perceive as meaningful, distinguishing clinically relevant changes from statistically significant but trivially small differences. Without the MID, it is impossible to determine whether an observed change in FACE-Q score after surgery represents a meaningful improvement, a meaningful deterioration, or merely measurement noise.

The MID serves multiple purposes. For individual patient care, it lets clinicians to interpret serial FACE-Q scores and counsel patients about whether a change is clinically meaningful. For research, it is needed for calculating sample sizes for clinical trials and for defining responder rates in comparative effectiveness studies. For quality improvement, it helps identify of patients with suboptimal outcomes. Regulatory agencies and professional guidelines increasingly require MID-based interpretation of PROM endpoints.

Two broad approaches to MID estimation exist. Anchor-based methods relate the change in PROM score to an external criterion (anchor) that captures the patient’s global perception of change. Distribution-based methods define the MID in terms of the statistical properties of the score distribution, such as a fraction of the standard deviation or the standard error of measurement. We combined results from both approaches, which is standard practice.

The aim of this study was to determine the MID for all four FACE-Q Skin Cancer Module scales using both distribution-based and anchor-based approaches, in a prospective cohort of patients undergoing facial skin cancer excision and reconstruction.

## METHODS

### Study Design and Participants

This was a prospective cohort study conducted at the Department of Plastic and Reconstructive Surgery, Catharina Ziekenhuis, Eindhoven, the Netherlands (2017–2018). Consecutive adult patients undergoing excision and reconstruction of histologically confirmed facial skin cancer were enrolled. The FACE-Q Skin Cancer Module was administered preoperatively and at 1 week, 3 months, and 1 year postoperatively. The study was approved by the institutional ethics committee.

### FACE-Q Skin Cancer Module

The module comprises four independently functioning scales. The Appearance Satisfaction scale (9 items, response range 1–5, sum score 9–45) measures satisfaction with facial appearance, with higher scores indicating greater satisfaction. The Psychosocial Distress scale (8 items, range 1–5, sum score 8–40) captures appearance-related psychological distress, with higher scores indicating more distress. The Cancer Worry scale (10 items, range 1–5, sum score 10–50) assesses worry about cancer recurrence. The Scar Satisfaction scale (8 items, range 1–5, sum score 8–40) measures satisfaction with surgical scars, available only postoperatively. All sum scores can be linearly transformed to a 0–100 scale.

### Distribution-Based MID Methods

Three distribution-based approaches were used. First, the 0.5 standard deviation (SD) method, based on the finding by Norman et al. that MIDs for quality-of-life instruments typically approximate half a standard deviation of baseline scores. Second, the standard error of measurement (SEM), calculated as SD × √(1 − α), where α is Cronbach’s alpha as a measure of internal consistency reliability. The SEM represents the precision of measurement and has been shown to correspond closely to the MID. Third, the 0.2 SD and 0.3 SD were calculated as sensitivity bounds representing the lower range of potentially meaningful change.

### Anchor-Based MID Method

The FACE-Q Adverse Effects scale, which measures treatment-related symptoms (pain, discomfort, numbness, etc.; 10 items, sum score 10–40), was used as an implicit anchor. This scale is administered at 1 week and 3 months postoperatively, and a decrease in effects score indicates symptomatic improvement. Patients were categorized into five groups based on their change in effects score from 1 week to 3 months: “much improved” (decrease >5 points), “somewhat improved” (decrease 1–5 points), “stable” (change ±1 point), “somewhat worsened” (increase 1–5), and “much worsened” (increase >5). The MID was defined as the mean change in FACE-Q score in the “somewhat improved” group, consistent with the standard anchor-based methodology.

### Statistical Analysis

Internal consistency was assessed using Cronbach’s alpha for each scale at each timepoint. Change scores were calculated for all available paired timepoints and compared using paired t-tests. Responsiveness was quantified using Cohen’s d (mean change / SD of baseline) and the standardized response mean (SRM; mean change / SD of change). Floor and ceiling effects were defined as >15% of respondents scoring the minimum or maximum possible score. All analyses were performed in Python 3.10.

The final recommended MID for each scale was arrived at by averaging the two methods, taking the mean of the 0.5 SD and SEM estimates and rounding to the nearest 0.5 sum points.

## RESULTS

### Study Population and Data Availability

A total of 287 patients were enrolled. Response rates were 252 (87.8%) at 1 week, 220 (76.7%) at 3 months, and 168 (58.5%) at 1 year. After matching on patient ID, 111 patients had paired preoperative–three-month data for the Appearance and Psychosocial scales, 111 for Cancer Worry, and 106 had paired 1-week–three-month data for Scars.

### Descriptive Statistics

Table 1 presents descriptive statistics for all scales across timepoints. Mean baseline Appearance Satisfaction was 29.0 ± 5.1 (sum score), Psychosocial Distress was 12.6 ± 3.9, and Cancer Worry was 20.1 ± 5.9. No significant floor or ceiling effects were observed (all <7%). All scales had strong internal consistency: Appearance α = 0.93, Psychosocial α = 0.82, Cancer Worry α = 0.90, and Scars α = 0.93.

### Change Scores and Responsiveness

Table 2 presents change scores across timepoints. The Cancer Worry scale showed the largest and most significant improvement from baseline to three months (mean change −3.14 ± 5.80; t = −5.70; p < 0.001; SRM = −0.54), representing a medium effect size. Scar satisfaction improved significantly from 1 week to 3 months (+3.02 ± 5.55; SRM = 0.54; p < 0.001). Appearance Satisfaction showed a small, non-significant improvement from baseline to 3 months (+0.59 ± 5.70; SRM = 0.10; p = 0.27), but significant improvement from 1 week to 3 months (+1.98 ± 6.89; SRM = 0.29; p = 0.004). Psychosocial Distress increased marginally from baseline to 3 months (+1.18 ± 4.88; SRM = 0.24; p = 0.01), suggesting slightly more distress at follow-up.

At the individual level, 54.1% of patients showed improved Appearance scores at 3 months, while 34.2% showed deterioration. For Cancer Worry, 63.1% improved and 22.5% worsened.

### Distribution-Based MID Estimates

Table 3 presents the distribution-based MID estimates. The 0.5 SD estimates were: Appearance 2.5, Psychosocial 2.0, Cancer Worry 3.0, and Scars 2.4 (sum score points). The SEM estimates were: Appearance 1.4, Psychosocial 1.7, Cancer Worry 1.9, and Scars 1.3. The SEM was consistently smaller than the 0.5 SD, as expected given the high reliability of all scales.

**Table 3.**
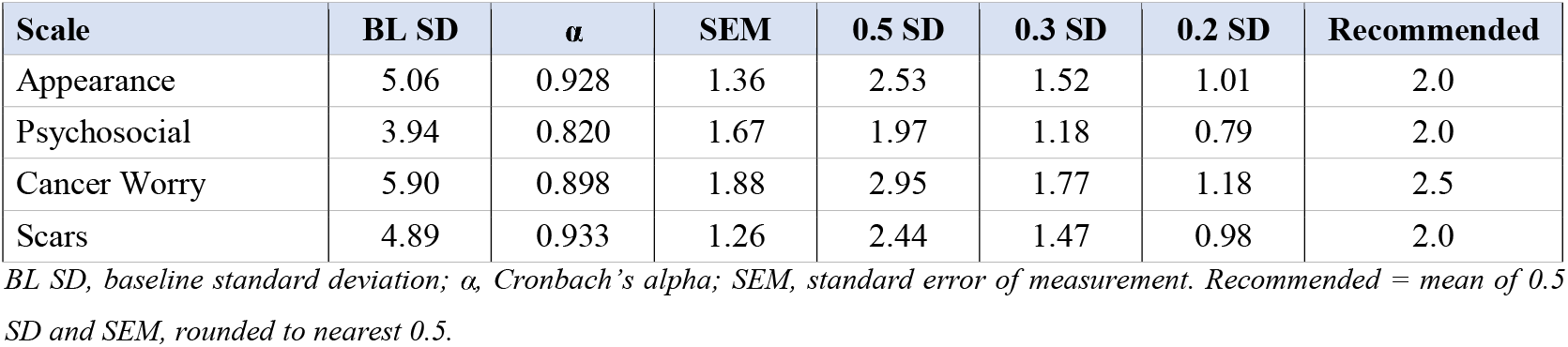
Distribution-based MID estimates for FACE-Q Skin Cancer Module scales.

### Anchor-Based MID Estimates

Table 4 presents anchor-based results using the Adverse Effects scale. Of 106 patients with paired 1-week–3-month data, 41 (38.7%) showed much improvement in adverse effects, 45 (42.5%) showed some improvement, 12 (11.3%) were stable, and 8 (7.5%) worsened. For the Scar scale, the anchor-based MID (mean change in the “somewhat improved” group) was 2.4 sum points, closely aligning with the distribution-based estimates. For Appearance, the anchor-based estimate was 1.0, lower than distribution-based estimates, possibly reflecting the indirect nature of this anchor.

### Triangulated MID Recommendations

Combining distribution-based and anchor-based estimates, the recommended MID values are presented in Table 5. On the sum score scale, the MID is 2.0 points for Appearance Satisfaction, 2.0 for Psychosocial Distress, 2.5 for Cancer Worry, and 2.0 for Scar Satisfaction. Converted to the 0–100 scale, these correspond to approximately 5.6, 6.2, 6.2, and 6.2 points, respectively. All four scales fall within a narrow range of 5–7 points on the 0–100 scale, matching what others have found on quality-of-life instruments.

**Table 5.** Recommended MID values for the FACE-Q Skin Cancer Module.

| Scale | MID (sum) | MID (0–100) | Interpretation |
| --- | --- | --- | --- |
| Appearance | 2.0 | 5.6 | A 2-point improvement in sum score is clinically meaningful |
| Psychosocial | 2.0 | 6.2 | A 2-point reduction in distress score is clinically meaningful |
| Cancer Worry | 2.5 | 6.2 | A 2.5-point reduction in worry is clinically meaningful |
| Scars | 2.0 | 6.2 | A 2-point improvement in scar score is clinically meaningful |

## DISCUSSION

This study establishes the first MID values for the FACE-Q Skin Cancer Module, filling a gap the validation studies left open. The triangulated MID of approximately 2–2.5 sum points (5–6 points on a 0–100 scale) across all four scales provides clinicians and researchers with actionable thresholds for interpreting individual score changes and designing clinical trials.

The fact that our MID estimates clustered together across scales (all within 5–7 points on the 0–100 scale) is consistent with the well-documented finding by Norman et al. that MIDs for quality-of-life instruments typically cluster around half a standard deviation, corresponding to 5– 8 points on a 0–100 scale. This agreement makes us more confident in our numbers and suggests that the FACE-Q Skin Cancer Module works like other well-tested patient questionnaires.

The responsiveness analysis provided important insights beyond the MID itself. Cancer Worry showed a medium effect size (SRM = −0.54), indicating that the FACE-Q is sensitive to the psychological relief patients experience after definitive excision of their cancer. Scar satisfaction improved significantly between 1 week and 3 months (SRM = 0.54), reflecting scar maturation during this period. In contrast, Appearance Satisfaction showed only a trivial change from baseline (SRM = 0.10), suggesting that reconstruction generally restores pre-cancer appearance levels rather than improving them—a finding with clear implications for patient counseling.

The marginal increase in Psychosocial Distress from baseline to 3 months is worth a closer look. While statistically significant, the mean change (+1.18 points) is below the MID, suggesting this increase is not clinically meaningful at the group level. Still, 40.5% of individual patients showed worsened psychosocial scores, showing that surgery affects people very differently to facial surgery.

Our study had some limitations. For one, the absence of an explicit patient-completed global rating of change (the gold standard anchor) meant we had to use the Adverse Effects scale as an implicit anchor. While this approach has precedent in the MID literature, the effects scale measures a different construct (physical symptoms) than the appearance and psychosocial scales, which may explain the lower anchor-based estimates for those scales. Future studies should prospectively collect a transition question. The distribution-based methods are independent of any anchor but may overestimate or underestimate the MID depending on the population’s baseline variability. And the SEM approach assumes that Cronbach’s alpha adequately represents test-retest reliability; ideally, stability data from patients with stable conditions would be used.

The practical implications of these MID values are substantial. In clinical practice, a clinician administering the FACE-Q before and after surgery can now interpret a change of ≥6 points on the 0–100 scale (or 2–3 sum points) as clinically meaningful. For trial design, the MID enables sample size calculations: a trial powered to detect the MID as a between-group difference with α = 0.05 and power = 0.80 would require approximately 80–100 patients per arm for the Appearance scale (assuming SD = 5.1) or 50–60 per arm for Cancer Worry (SD = 5.9).

### Limitations

This study has several limitations. First, the absence of a prospective global rating of change limits the anchor-based analysis. Second, the sample size of 111 paired observations, while adequate for distribution-based methods, limits the precision of subgroup-specific MID estimates. Third, the MID was established in a single-center Dutch population; cross-cultural validation is warranted. Fourth, these MID values apply to group-level interpretation; individual-level interpretation requires consideration of measurement error.

## CONCLUSIONS

This study establishes the MID for all four FACE-Q Skin Cancer Module scales at approximately 2–2.5 sum points (5–6 points on the 0–100 scale). These thresholds enable meaningful interpretation of individual score changes, responder analysis in clinical trials, and sample size calculations for future studies. The FACE-Q Skin Cancer Module demonstrates adequate responsiveness to postoperative change, particularly for cancer worry and scar outcomes. We recommend that future studies prospectively include a global rating of change to refine these estimates.

## Data Availability

The data underlying this study were collected at Clinique Rebelle, Amsterdam. Requests may be directed to the corresponding author. Clinic information: https://www.cliniquerebelle.com

## DECLARATIONS

### Ethical approval

Approved by the Medical Ethics Committee of Catharina Ziekenhuis (protocol [XXX]).

